# Physician Workforce and Population Mortality: A Globally Validated Age-Structured Modeling Approach

**DOI:** 10.1101/2025.06.16.25329730

**Authors:** Youngmin Oh, Eunju Lee

## Abstract

**Background:** Evaluating long-term health workforce planning requires robust and globally valid indicators. While many existing metrics, such as patient satisfaction or healthcare utilization, are inherently uncertain over long horizons, mortality is an objective and stable metric. To analyze how physician supply impacts population health, we propose a model that links physician density to age-specific mortality rates and incorporate it to the age-sturctured dynamics.

**Methods:** We developed a unified model that links physician workforce density to age-structured mortality rates. Subsequently, we applied this model within a Lotka-McKendrick framework to simulate physician supply expansion scenarios in South Korea. The system simulates future age-specific mortality outcomes under varying physician supply scenarios, including a baseline intake of 3,058 physicians per year and an expanded intake of up to 7,058 physicians per year, projected through 2065.

**Findings:** Our model was validated using WHO mortality data including Japan, the United States, and the United Kingdom. Its validity of our model holds across all age groups in each country, as confirmed by statistical analysis with false discovery rate correction (maximum adjusted *p <* 0.05). Using the age-structured dynamics with the model in South Korea, we confirm that future physician density increases even under the baseline scenario (3,058 physicians per year), and that projected population sizes under the baseline and the aggressive expansion scenario (7,058 physicians per year) are not statistically different (*p >* 0.07). Moreover, under the aggressive expansion scenario, the projected reductions in age-specific mortality rates by 2065 remain marginal: less than 0.27% for those under 65, less than 0.69% for ages 65–75, less than 2.75% for ages 75–84, less than 7.31% for ages 85–94, and less than 12.6% for ages 95–99.

**Interpretation:** In well-resourced health systems facing aging populations and persistently low fertility rates, further expansion of physician supply alone offers limited mortality benefits. Our findings suggest a paradigm shift: from quantity-driven to efficiency-focused workforce strategies. The proposed method is readily adaptable to other contries, offering a policy-relevant and outcome-oriented tool for long-term health workforce planning.

**Funding:** None. The funder had no role in study design, data collection, data analysis, data interpretation, or writing of the report.

**Research in context:** *Evidence before this study:* We searched PubMed, Web of Science, and Google Scholar for studies published between 2000 and 2024 using terms such as “physician workforce,” “healthcare workforce planning,” “medical workforce density,” and “age-structured dynamics.” Most physician workforce planning models are either utilization-based (projected service demand) or needs-based (estimated population health requirements). These models often extrapolate current health service utilization rates or population health needs into the future, typically assuming that such values remain constant over time, despite the acknowledged difficulty of forecasting long-term demand dynamics. Additionally, methodologies differ widely across countries, which hampers direct comparisons of outcomes. Some metrics, such as patient needs or preferences, are also subjective and difficult to measure consistently. In contrast, more objective indicators like mortality rates are more readily available and comparable across settings. As far as we are aware, no published model has dynamically integrated physician density—endogenously derived from workforce supply—into an age-structured framework for projecting population mortality.

*Added value of this study:* To the best of our knowledge, this study is the first to integrate a model linking physician workforce density with age-structured mortality rates into the Lotka–McKendrick framework, capturing the interaction between physicians and populations across age groups. By modeling how changes in doctor-to-population ratios affect mortality, our approach moves beyond traditional models that treat health outcomes as exogenous. This allows novel analyses of policy scenarios, such as estimating life expectancy gains from increasing physician supply. Our model provides a unified framework to project both physician workforce dynamics and population health outcomes simultaneously.

*Implications of all available evidence:* Our findings underscore the importance of explicitly linking age-structured population dynamics with physician density and mortality outcomes. Traditional models often overlook these interdependencies, risking inaccurate forecasts and limiting the relevance of cross-national comparisons. By capturing how physician supply influences mortality across age groups, our approach enables more precise, outcome-driven workforce planning.

## 1. Background

Developing a well-structured physician workforce planning is essential for an efficient healthcare system ^1^. A shortage of physicians limits access to medical services, preventing patients from receiving necessary surgeries or treatments. In contrast, an oversupply of physicians leads to economic inefficiency, since physician education is time-consuming, labor-intensive, and entails substantial financial investment. To determine the optimal physician workforce size, various evaluation strategies for workforce planning have been proposed, broadly classified into needs-based, demand-based, supply projection, and benchmarking methods ^2–10^.

However, each approach has limitations. Needs-based methods rely on subjective assessments and may overlook patients who need medical services but do not recognize their own needs ^11,12^. Demand-based methods, which project future need from past utilization, can over-or underestimate physician requirements—especially when demographic ageing, insurance-coverage expansions, or other structural changes are mis-specified—thereby risking both over-and undersupply ^13,14^. Supply projection methods only predict future physician numbers without determining the required workforce. Consequently, conducting a post-hoc analysis is ultimately indispensable ^15^. Benchmarking, based on reference countries, assumes cross-system comparability, which may not hold ^16^. In particular, these methods often rely on factors that are difficult to measure and predict, such as physician productivity, technological advancements, and working hours ^16–18^.

To address these challenges, we propose a simple yet robust alternative based on Occam’s razor principle ^19^. A key factor of our approach is developing a model linking physician density to age-structured mortality rates. Ambiguous measures such as patient satisfaction or quality-of-life indicators are difficult to quantify and forecast over long periods ^20^. In contrast, these mortality rates have been historically and universally measured in most developed countries. Being statistically stable and less sensitive to national policy fluctuations, mortality rates serve as one of the most reliable features for assessing long-term health system performance ^21–23^.

We evaluate the model across multiple countries using WHO data ^24^ and confirmed its statistical validity: the largest F-test adjusted *p*-value remained below 0.05 after false-discovery-rate (FDR) correction at *α* = 0.005. This cross-national validation positions our framework as a globally applicable tool for workforce-policy evaluation rather than a country-specific model. We then embed the validated model within South Korea’s age-structured demographic dynamics ^25,26^, integrating age-specific fertility, population, and mortality data from KOSIS.

Note that only basic demographic and workforce data are required to construct our model, making it highly tractable and transparent. By integrating this model with a mathematical framework for population dynamics—the Lotka–McKendrick model ^25,26^—we enable long-term scenario simulations of age-structured mortality rates and physician density. These simulations offer policymakers a reliable evidence base for evaluating the population-level impacts of various workforce strategies.

## 2. Methods

Let *t* denote time and *a* denote age. We define the population and physician workforce as *p*(*t, a*) and *w*(*t, a*), respectively. The corresponding age-specific mortality rates are given by ℳ*_p_*(*t, a*) for the general population and ℳ*_w_*(*t, a*) for the physician workforce. The initial age distributions are denoted by *p*_0_(*a*) and *w*_0_(*a*), respectively. We also define ℳ*_t_*(*t, a*) as the age-specific fertility rate at time *t*, and *m_t_* as the medical workforce supply plan at time *t*.

A high population relative to physicians may prevent all patients from taking healthcare services sufficiently, while a larger workforce expands healthcare coverage but increases financial burdens to educate and maintain the experts. Following this observation, we model age-structured mortality rates ℳ*_p_*(*t, a*) as a function of physician workforce density *ρ*(*t*):

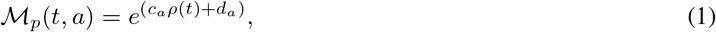

where

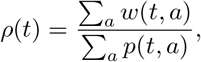

with *c_a_* and *d_a_* as age-dependent but time-independent coefficients. Eq. (1) implicitly assumes the law of large numbers ^27^, which holds when the physician workforce is sufficiently large and medical advancements are not considered. We assume that all physicians supplied have the same age *a* = 26. Moreover, since physicians generally have shorter lifespans and many continue working beyond age 65, we make the conservative assumption that all physicians retire at *a*^*^ ≥ 65 and that those younger than 65 have the same mortality rates as the general population. Although the official retirement age for medical professors in South Korea is 65, setting this age as a fixed threshold can be considered a conservative assumption in light of current medical workforce dynamics. In practice, many physicians continue clinical work beyond the age of 65, either through reappointment at university hospitals or by transitioning to private clinics and smaller healthcare institutions ^28–33^. Finally, we assume that the effect of migration on the total population is negligible. In summary, our analysis is based on the following assumptions:

### Assumption 1.

*The individual abilities of each physician have minimal impact on age-structured mortality rates.*

### Assumption 2.

*The effect of medical advancements on mortality reduction is not considered.*

### Assumption 3.

*All physicians supplied have the same age a* = 26.

### Assumption 4.

*The mortality rates between the population and physicians are the same.* ℳ*_w_*(*t, a*) = ℳ*_p_*(*t, a*).

### Assumption 5.

*Future total populations are independent of international migration inflows.*

### Assumption 6.

*All physicians retire at the same age threshold a*^*^ = 65, *i.e.,*

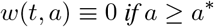

*for all t.*

Now, combining the Lotka-McKendrick system ^25^ with Eq. (1) under the assumptions, we obtain the dynamics of the population and physician workforce: For the population *p*(*t, a*),

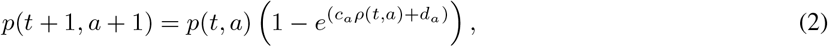

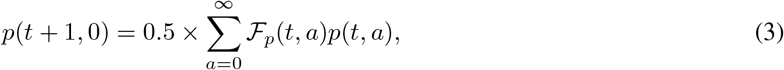

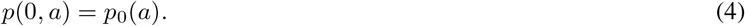

For the workforce,

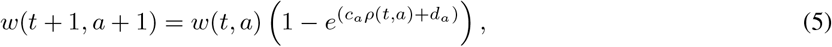

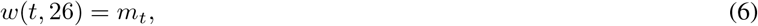

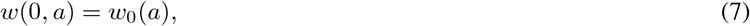

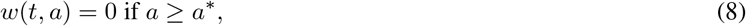

Eq. (6)–Eq. (7) follow from assumption 3 and assumption 4, respectively, and Eq. (8) follows from assumption 6. One can observe that the systems (2)–(4) and (5)–(7) interact through *ρ*(*t, a*), which represents physician density and affects the mortality rates as described in Eq. (1). The detailed derivations are provided in the suplementary material.

## 3. Results

### 3.1. Validation of the Modeling ℳ*_p_* (Eq. (1))

Figure 1 demonstrates the model’s robustness across countries in the WHO dataset ^24^. It can be observed that linear regressions fit well across age groups in different countries. The FDR-corrected *p*-values from F-tests at a significance level of *α* = 0.05 were less than 0.05 across all age-groups for each country. These consistently significant results support the validity of the model across heterogeneous populations and countries. In South Korea, we validated Eq. (1) using national statistics from 2006–2023. To achieve a more granular analysis, we subdivided the age groups more finely. Linear regression fits of log-mortality to physician density for each Korean age group passed the F-test (highest FDR-adjusted *p*-value = 0.025 at *α* = 0.05), confirming validity across all age groups ^34^ (see Figure 1). Note that the age groups with *a <* 100 have negative slopes, indicating that higher physician density is associated with lower mortality rates. For individuals with *a <* 100, the 95% confidence intervals for the slopes were strictly negative (see Figure 2). The positive slope observed in the 100+ age group reflects cumulative survival gains at earlier ages, which result in an increased proportion of individuals reaching extreme old age, where mortality is inevitably higher. Consequently, as more individuals survive into extreme old age, mortality becomes concentrated in the 100+ age group.

**Figure 1.**
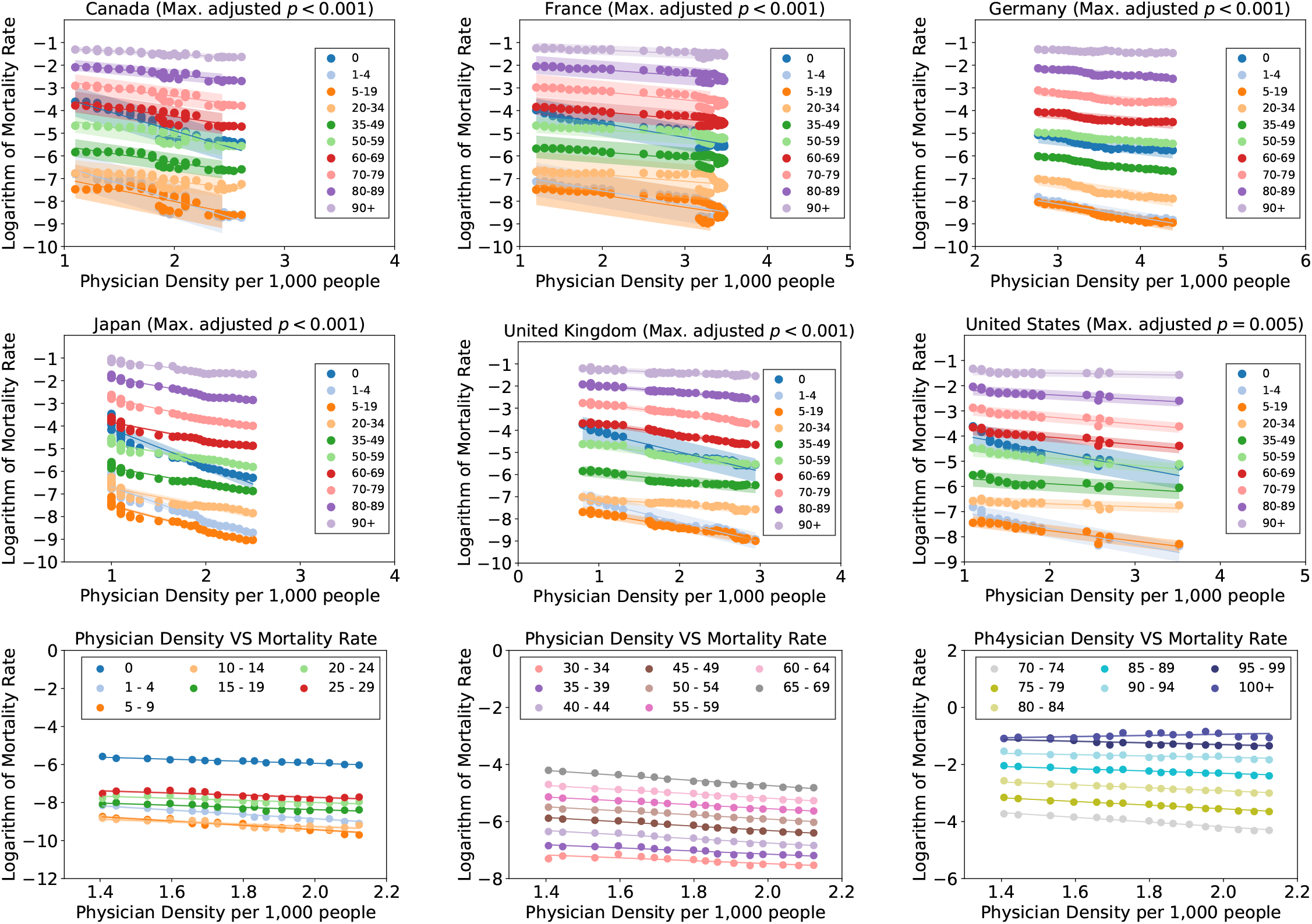
Linear regression of logarithmic mortality rate against physician density (1980-2022) across various contries. The highest adjusted *p*-value across all age groups in F-tests was smaller than 0.05 after flase discovery rate correction. The solid line represents the mean, with shaded regions indicating 95% confidence intervals.

**Figure 2.**
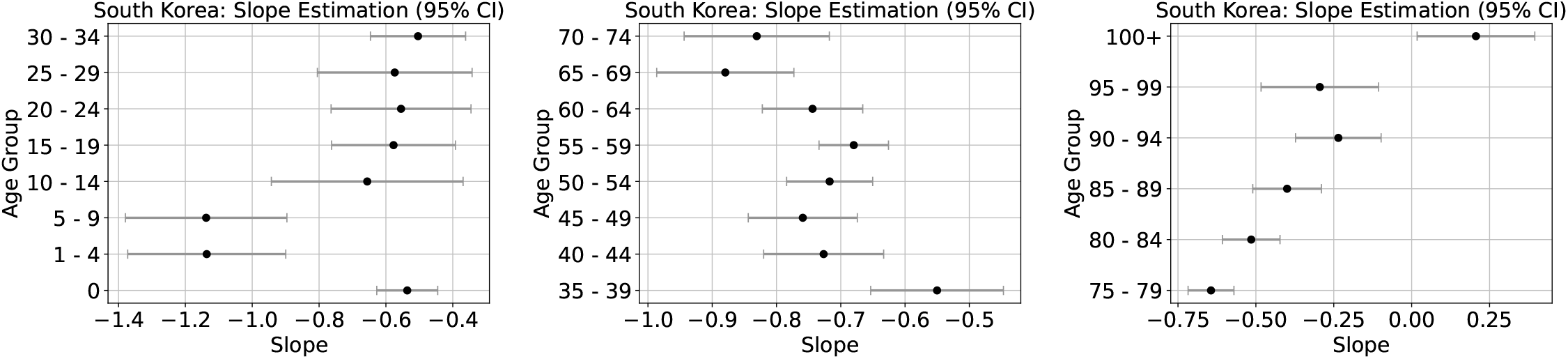
Estimation of slopes for each age group under 95% confidence bound in South Korea. Notably, the slope for individuals aged 100 and above is positive, in contrast to the negative slopes observed in all other age groups.

### 3.2. Prediction via Age-structured Dynamics

#### 3.2.1. Physician Workforce Planning

We consider the following four annual physician supply planning strategies to evaluate the impact of the expansion:

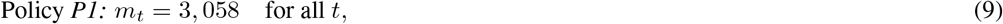

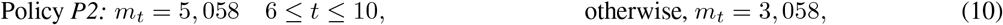

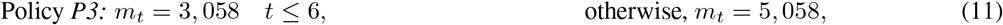

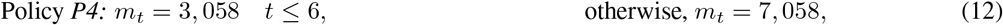

The planning *P1* in Eq. (9) represents the annual physician supply, which corresponds to South Korea’s medical school admission quotas through 2024. *P2*-*P4* in Eq.(10)-(12) propose expansions of annual physician supply.

#### 3.2.2. Specifying Components in the Governing Equations

We determine initial conditions for *p*_0_(*a*) and *w*_0_(*a*). We use the population data in the year 2024 published in KOSIS as the initial condition *p*_0_(*a*). In the case of *w*(0, *a*), there are 2.2 physicians per 1,000 population in South Korea ^35^ in the year 2024. Thus, we use the physician age ratio from ^36^ and distribute the physician population:

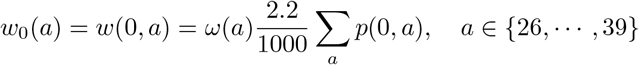

where

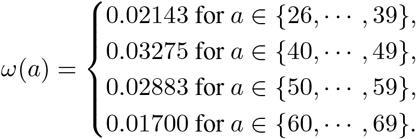

Note that we assume that physicians who is older than 65 retired without any exception, i.e., *w*(*t, a*) ≡ 0 for *a ≥ a*^*^ = 65 and *t ≥* 0.

Now, we determine the fertility rates ℱ*_p_*(*t, a*). These fertility rates are influenced by various factors, including housing policies, making their prediction uncertain. To establish a robust upper bound, we use the age-structured fertility rates from 2007, when the total fertility rate was 1.25 because the fertility has declined across most age groups ^37^ in South Korea. Besides, projections by KOSIS estimate the rates will remain below 1.07 until 2065^38^. Therefore, if we can conclude that the number of physicians in sufficienty under the 2007 fertility data, our conclusions remain robust.

In detail, ℱ*_p_*(*t, a*) can be formulated as

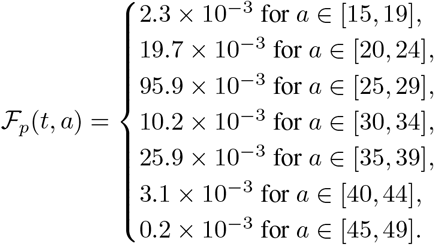

#### 3.2.3. Predicting Future Population and Physician Workforce

We analyze four physician supply planning scenarios (*P1*-*P4*). Figure 3.(a)-(d) present the projected population trends through 2065, showing minimal differences across scenarios. The Mann-Whitney U test yields *p*-values of 0.623, 0.268, and 0.073 for *P1* compared to *P2, P3*, and *P4*, respectively, confirming the similarity in population projections. In contrast, Figure 3.(e)-(h) illustrate the projected physician workforce over different planning scenarios, revealing significant disparities across scenarios (*p <* 0.001 for *P1* vs. all others). Notably, the temporal reductions in physicians appear in Figure 3.(e)-(f) as a result of the impact of physician retirements, but not in Figure 3.(g)-(h) due to the high physician supplements. Despite these differences, Figure 3.(i)-(l) demonstrates that physician density increases across all scenarios, independent of policy differences. Linear regression on physician density trends confirms this, with all F-test *p*-values below 0.001 and consistently positive slopes (with 99% confidence bound) across *P1*-*P4*. Thus, the baseline scenario (*P1*) even ensures a steady rise in physician density with high probability.

**Figure 3.**
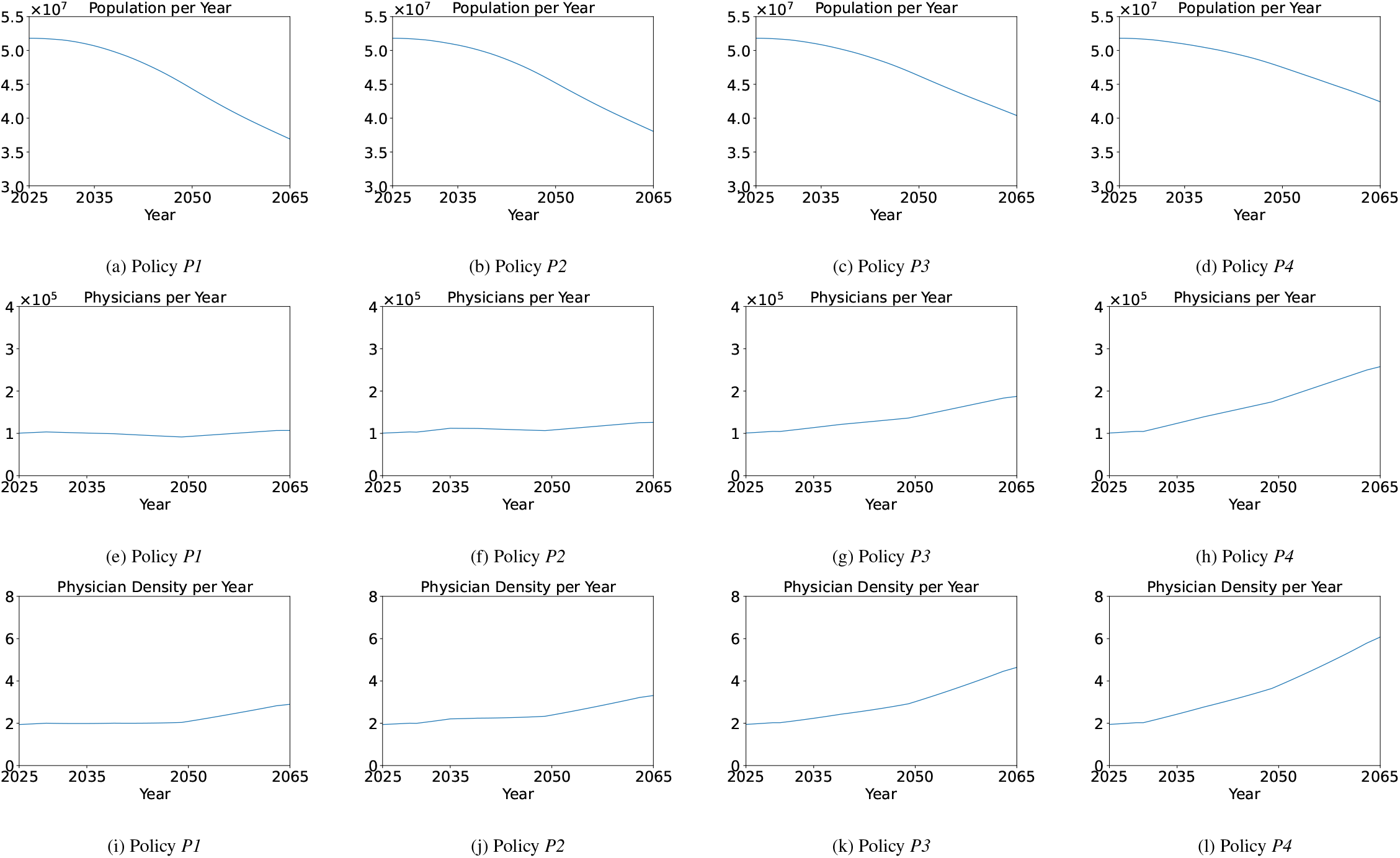
(a)-(d): Predicted population curves under each supply strategy. Mann-Whitney U test showed no significant differences between *P1* (9) and *P2* (10), *P3* (11), *P4* (12) (*p* = 0.651, 0.303, 0.092). (e)-(h): Predicted physicians. Mann-Whitney U test confirmed significant differences (*p <* 0.001). (i)-(l): Predicted physician-per-1,000 ratio. F-tests for each linear regression were valid (*p <* 0.001). Moreover, Linear regression slopes were positive under 95% confidence bound.

The complete source code used for simulation and regression analysis is publicly available at: https://anonymous.4open.science/r/medical_quota-2756

### 3.3. Predicting Future Mortality Rates

Figure 4 shows projected mortality rates under physician supply scenarios *P1*–*P4* (Eqs. (9)–(12)). Mortality decreases for *a <* 100 but rises for *a ≥* 100 due to improved healthcare access. Table 1 presents expected mortality reductions per 100,000 by the year 2065 compared to *P1*. The absolute mortality reduction remains *<* 0.24% for those under 65, *<* 1.26% for ages 65–79, and *<* 11.9% for those ≥ 80, even between 3,058 or 7,058 annual physician supplies. While additional physician supply leads to some mortality reduction among the older population, the magnitude of improvement is modest relative to the substantial economic costs associated with expanding the workforce to 7,058 per year. This suggests that further expansion of physician supply may produce limited health gains, particularly in already well-resourced systems.

**Table 1.** Expected Mortality Reduction (%) per 100,000 people compared to the original physician supply planning *P1*.

| Age Group | *P2* (%) | *P3* (%) | *P4* (%) | Age Group | *P2* (%) | *P3* (%) | *P4* (%) |
| --- | --- | --- | --- | --- | --- | --- | --- |
| 0 | 0.03242 | 0.09898 | 0.13420 | 50–54 | 0.03674 | 0.10238 | 0.12974 |
| 01–04 | 0.00198 | 0.00459 | 0.00521 | 55–59 | 0.05216 | 0.14806 | 0.19004 |
| 05–09 | 0.00111 | 0.00258 | 0.00293 | 60–64 | 0.07707 | 0.21207 | 0.26638 |
| 10–14 | 0.00125 | 0.00358 | 0.00463 | 65–69 | 0.12319 | 0.31822 | 0.38385 |
| 15–19 | 0.00292 | 0.00873 | 0.01165 | 70–74 | 0.21228 | 0.56071 | 0.68576 |
| 20–24 | 0.00419 | 0.01267 | 0.01705 | 75–79 | 0.37653 | 1.08799 | 1.41447 |
| 25–29 | 0.00556 | 0.01666 | 0.02225 | 80–84 | 0.65013 | 2.00746 | 2.74603 |
| 30–34 | 0.00677 | 0.02102 | 0.02889 | 85–89 | 1.05122 | 3.45405 | 4.97632 |
| 35–39 | 0.00981 | 0.02975 | 0.04010 | 90–94 | 1.29037 | 4.65710 | 7.30954 |
| 40–44 | 0.01592 | 0.04416 | 0.05579 | 95–99 | 2.37052 | 8.26441 | 12.55571 |
| 45–49 | 0.02493 | 0.06810 | 0.08512 | 100+ | -4.07084 | -19.47555 | -41.33239 |

**Figure 4.**
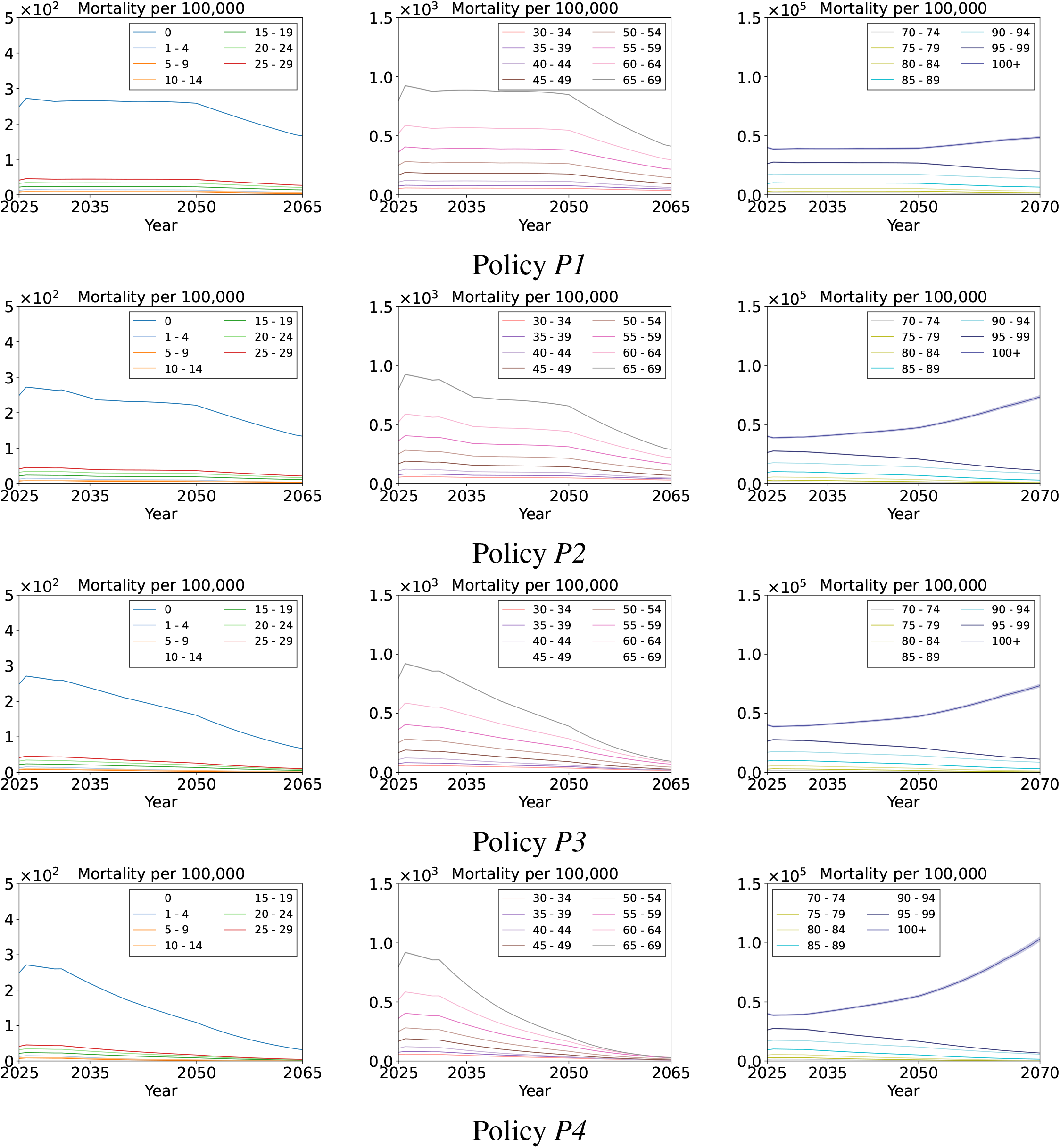
The predicted mortality rates per each age group under (a) *P1*: annual physician supplies 3,058, (b) *P2*: annual physician supplies 3,058 with an increase to 5,058 for the first five years, (c) *P3*: annual physician supplies 5,058, and (d) *P4*: annual physician supplies 7,058.

We additionally provide linear regression analyses for other countries using WHO data to further validate our model (Eq. 1) in the supplementary material. Furthermore, sensitivity analyses on physicians’ retirement age, fertility rates, and reductions in physician supply are provided, respectively, in the supplementary material.

## 4. Discussion

We proposed an age-structured population model linking physician density to age-structured mortality rates. The model was validated using WHO data from various countries through linear regression, yielding adjusted *p*-values below 0.05 after false discovery rate correction at *α* = 0.05. By integrating the model with the Lotka–McKendrick system and South Korea’s demographic data, we simulated various physician supply scenarios and quantified their impact on age-specific mortality rates through 2065.

In South Korea—where physician accessibility is among the highest and mortality rates among the lowest in OECD countries—even a twofold expansion of the physician workforce resulted in only marginal reductions in mortality. This result held consistently under conservative assumptions such as universal physician retirement at age 65 and the exclusion of future medical advancements. These findings suggest that further expanding the physician supply alone is unlikely to yield substantial improvements in mortality outcomes or overall population health indicators.

From a health policy perspective, these results carry important implications. Expanding the physician workforce requires considerable economic resources due to the long training periods, infrastructure needs, and ongoing costs. When the marginal benefit to health outcomes is limited, the cost-effectiveness of supply-side expansion becomes questionable. Policymakers must therefore critically assess whether such strategies represent the most efficient use of health system resources.

Instead, alternative approaches prioritizing redistributive efficiency and targeted deployment may yield greater returns. In the Korean context, notable disparities persist in physician distribution between urban centers and rural or aging regions. A more targeted policy mix may therefore offer higher marginal benefits to public health. These include incentive-based rural placement programs, scholarship-linked service in underserved areas, expanded residency and training opportunities in regional hospitals. Such strategies are likely to provide more sustainable and equitable improvements in healthcare access than undifferentiated workforce expansion.

More broadly, the model’s minimal data requirements—age-structured mortality and physician density—make it applicable beyond the Korean setting. In particular, it is well-suited for adaptation in other high-income countries facing population aging, such as Japan, Germany, or the United Kingdom. These nations share demographic and workforce characteristics such as low fertility rates, high physician densities, and mature healthcare systems. For such countries, the model offers a generalizable, outcome-oriented tool for evaluating diminishing returns in workforce expansion and for informing long-term planning. Future extensions could incorporate morbidity indicators or access-based health metrics to further strengthen its cross-national applicability.

In conclusion, while maintaining adequate physician supply remains fundamental, indiscriminate expansion is unlikely to meaningfully enhance health outcomes in already well-resourced systems. A more nuanced, multi-pronged strategy emphasizing redistributive allocation and efficiency-enhancing innovations presents a more sustainable path toward resilient healthcare systems under demographic pressure.

### 4.1. Limitations

Our model is based on several simplifying assumptions that merit discussion. First, it assumes that physician supply is sufficiently large to allow individual-level variation in productivity, working hours, and quality to be averaged out at the system level This assumption is supported by the law of large numbers and is generally valid in countries with large and mature medical workforces such as Japan, United Kingdoms, and South Korea. However, in health systems with small or highly heterogeneous workforces, such as in some low-income or sparsely populated countries, this approximation may not hold. Second, the model assumes limited international physician migration. In socially stable countries with strong health governance—such as South Korea, Japan, or Germany—physician supply trends are largely determined by domestic training and retirement patterns. However, in countries facing political instability, economic crisis, or high rates of medical brain drain, physician density may fluctuate substantially due to external migration. In such settings, long-term workforce planning based on closed-population assumptions may lead to inaccurate or misleading projections. Third, we exclude medical technology progress and epidemiological transitions from the model. While these factors can influence age-specific mortality over time, they are extremely difficult to predict over multi-decade horizons. Given the lack of consistent long-term indicators and high uncertainty, their exclusion may be justified in favor of model tractability and stability. Lastly, our outcome variable is limited to mortality. Mortality is a robust, objective, and internationally comparable measure, which supports its use in long-term forecasting models. However, it does not capture subjective or functional aspects of health system performance—such as morbidity, patient satisfaction, or quality of life—which may be increasingly relevant in aging societies. Future research could extend this framework to include such metrics where reliable data allow.

## Data sharing

All data and code used for the simulations in this study are publicly available at https://anonymous.4open.science/r/medicalquota-2756. The shared materials include:

- Model source code and analysis scripts.
- Simulated data generated from public WHO and KOSIS databases; no individual participant data are included. Additional documents (study protocol and statistical analysis plan) are not applicable to this modeling study.

Data are available with publication, with no restrictions; access is open to all users via the provided URL.

## Data Availability

WHO, KOSIS

https://kosis.kr/index/index.do

https://data.who.int/

https://www.who.int/data/gho/

## Acknowledgments

This research did not receive any specific grant from funding agencies in the public, commercial, or not-for-profit sectors.

