## Supplemental Material for "Physician Workforce and Population Mortality: A Globally Validated Age-Structured Modeling Approach"

### Appendix A. Derivation of System in Eq. (2)–Eq. (7)

In this paper, we use the Lotka-McKendrick equation[26] for age-structured dynamics to predict a quantity population $N(t, a)$ having age till $a > 0$ at time $t$. $N(t, a)$ is modeled as

$$\begin{aligned} N(t+h, a+h) = N(t, a) + \frac{1}{2}\int_{t}^{t+h} \mathcal{F}(s) ds \\ - \int_{s=0}^{h}\int_{a=0}^{a+s} \mathcal{M}(t+s, \sigma) p(t+s, \sigma) d\sigma ds \end{aligned} \tag{A.1}$$

for infinitesimal $h$, the supply rate density function $\mathcal{F}(t, a)$, and the mortality rate density function $\mathcal{M}(t, a)$, respectively, where

$$N(t, a) = \int_{0}^{a} q(t, \sigma) d\sigma. \tag{A.2}$$

with a density function $q(t, a)$. Differentiating Eq. (A.1) on $h$ with $h = 0$ leads to

$$q(t, a) + \int_{0}^{a} q_t(t, \sigma) d\sigma = \frac{1}{2}\mathcal{F}(t) - \int_{0}^{a} \mathcal{M}(t, \sigma) q(t, \sigma) d\sigma. \tag{A.3}$$

Differentiating Eq. (A.3) on $a$ again, we obtain the following system:

$$q_a(t, a) + q_t(t, a) = \mathcal{M}(t, a) q(t, a), \tag{A.4}$$

$$q(t, 0) = \frac{1}{2}\int_{0}^{\infty} \mathcal{F}(t, \sigma) q(t, \sigma) d\sigma, \tag{A.5}$$

$$q(0, a) = q_0(a), \tag{A.6}$$

where Eq. (A.4) is the governing equation, Eq. (A.5) represents supported populations by $\mathcal{F}(t, a)$, and Eq. (A.6) is an initial condition. Note that Eq. (A.4)–Eq. (A.6) is the continuous-time system $t$. By substituting the following equations

$$\begin{aligned} q_a(t, a) &= \frac{q(t, a) - q(t, a-h)}{h}, \\ q_t(t, a) &= \frac{q(t, a-h) - q(t-h, a-h)}{h}, \end{aligned}$$

into Eq. (A.4), where $h$ is an infinitesimal, we can obtain the following system with $h = 1$:

$$q(t, a) = q(t-1, a-1)(1 - \mathcal{M}(t-1, a-1)), \tag{A.7}$$

$$q(t, 0) = \frac{1}{2}\sum_{a=0}^{\infty} \mathcal{F}(t-1, a) q(t-1, a), \tag{A.8}$$

$$q(0, a) = q_0(a), \tag{A.9}$$

for $t, a \in \mathbb{N}$, a set of natural numbers and the zero. In other words, the system Eq. (A.7)–Eq. (A.9) is a discrete variant of Eq. (A.4)-Eq. (A.6). To uniquely define the quantity $q(t, a)$ for all $t, a$, it is necessary to determine the crucial three components:

1. Initial condition: $q(0, a)$,
2. Supply rate on $t$ and $a$: $\mathcal{F}(t, a)$,

3. Mortality rate on $t$ and $a$: $\mathcal{M}(t, a)$.

Let $p(t, a)$ denote the population of age $a$ at time $t$, with mortality rate $\mathcal{M}_p$ and fertility rate $\mathcal{F}_p$. The population dynamics are governed by the following system:

$$p(t, a) = p(t-1, a-1)\left(1 - \mathcal{M}_p(t-1, a-1)\right), \tag{A.10}$$

$$p(t, 0) = \frac{1}{2}\sum_{a=0}^{\infty} \mathcal{F}_p(t-1, a)\, p(t-1, a), \tag{A.11}$$

$$p(0, a) = p_0(a), \tag{A.12}$$

Let $w(t, a)$ denote the physician workforce of age $a$ at time $t$, with mortality rate $\mathcal{M}_w$. In the case of $w(t, a)$,, we need to modify Eq. (A.8) by employing the quantity $m_t$ the number of newly certified physicians entering the workforce at time $t$, assuming that medical students typically obtain their certification at age $a = 26$ as

$$w(t+1, 26) = m_t, \tag{A.13}$$

$$w(t+1, a) = 0, \quad \text{for } a \neq 26. \tag{A.14}$$

By replaing Eq. (A.8) with Eqs. Eq. (A.13)-Eq. (A.14), we obtain

$$w(t+1, a+1) = w(t, a)\left(1 - \mathcal{M}_w(t-1, a-1)\right), \tag{A.15}$$

$$w(t+1, 26) = m_t, \tag{A.16}$$

$$w(t+1, a) = 0 \quad \text{for } a \neq 26, \tag{A.17}$$

$$w(0, a) = w_0(a). \tag{A.18}$$

Finally, by assuming $\mathcal{M}_w(t, a) = \mathcal{M}_p(t, a)$ and substituting $\mathcal{M}_p(t, a) = \mathcal{M}_w(t, a) = e^{(c_a \rho(t) + d_a)}$ into Eq. (A.10) and Eq. (A.17), we obtain the system Eq. (A.19)-Eq. (A.24) below in the end: For the population $p(t, a)$,

$$p(t+1, a+1) = p(t, a)\left(1 - e^{(c_a \rho(t,a) + d_a)}\right), \tag{A.19}$$

$$p(t+1, 0) = 0.5 \times \sum_{a=0}^{\infty} \mathcal{F}_p(t, a) p(t, a), \tag{A.20}$$

$$p(0, a) = p_0(a). \tag{A.21}$$

For the workforce,

$$w(t+1, a+1) = w(t, a)\left(1 - e^{(c_a \rho(t,a) + d_a)}\right), \tag{A.22}$$

$$w(t, 26) = m_t, \tag{A.23}$$

$$w(0, a) = w_0(a). \tag{A.24}$$

The two system: Eq. (A.19)–Eq. (A.24) are identical to the system explained in the main manuscript. Finally, we can impose a condition that all physicians retire at or after age $a^*$ such that

$$w(t, a) = 0 \quad \text{if } a \geq a^*.$$

### Appendix B. Additional Validness of Eq. (1)

To further validate our model in Eq. (1), we used additional countries from WHO data and analyzed the relationship between physician density and age-structured mortality rates. Figure Appendix B.1 and Figure Appendix B.2 illustrate the visual relationships across various countries. A consistent relationship can be observed in many cases. Adjusted $p$-values from F-tests, corrected for false discovery rate (FDR) at $\alpha = 0.05$, indicate that many of these countries are statistically governed by Eq. (1). These results demonstrate that our method is applicable to various countries, provided that fertility rates, initial population structures, and physician distributions are appropriately defined.

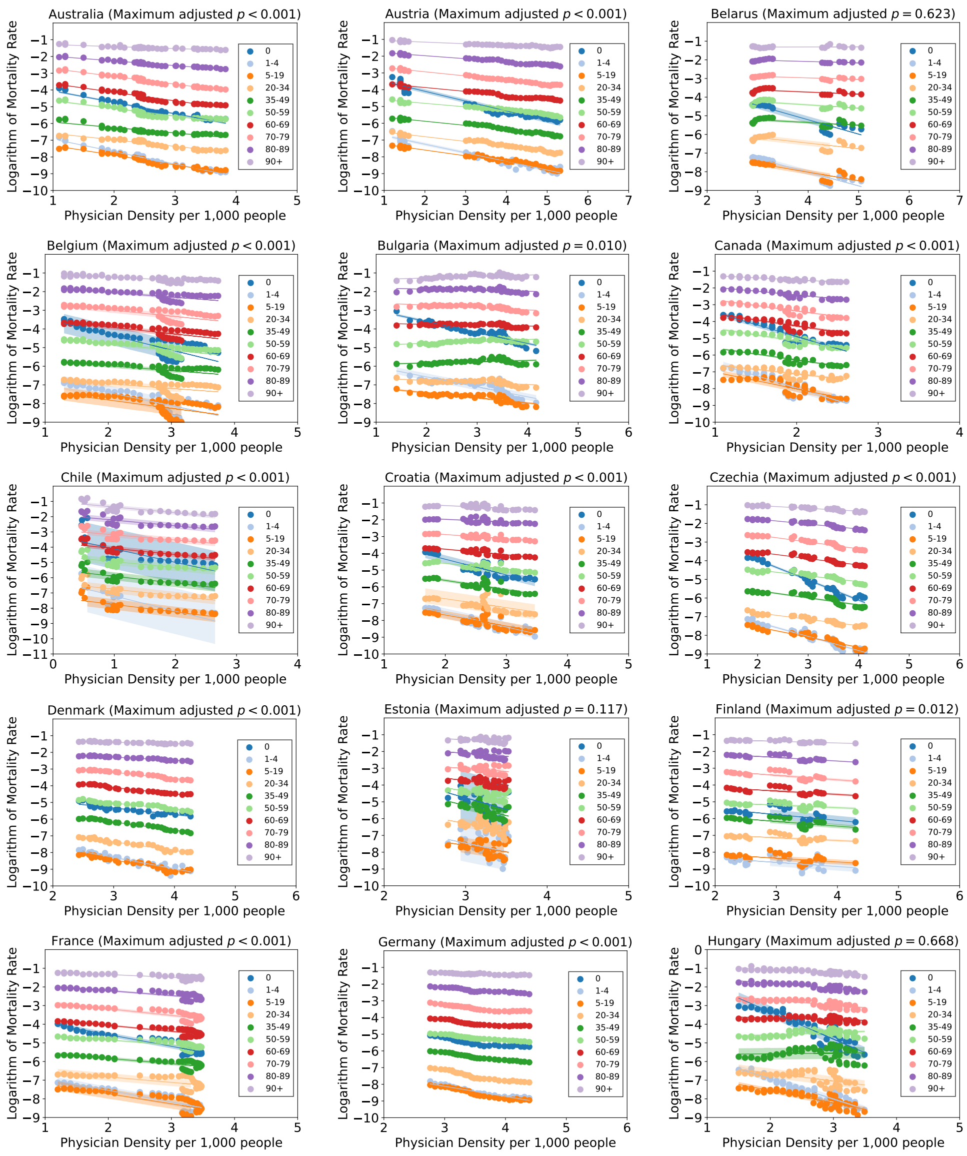


**Figure Appendix B.1:** Linear regression of logarithmic mortality rate against physician density (1960-2022) across all age groups. The solid line represents the mean, with shaded regions indicating 99.7% confidence intervals.

### Appendix C. Parameter Sensitivity Analysis

In this section, we examine the sensitivity of the age-structured dynamics discussed in the main manuscript. In the main analysis, we conservatively assumed that all physicians retire at age 65, given the uncertainty in forecasting future retirement patterns. In the current analysis, we vary this retirement age threshold to 70 and 75, respectively.

We conducted a thorough validation of the age-structured mortality rates under different retirement age thresholds to assess the consistency and robustness of the observed patterns. In particular, Figure Appendix C.1 and Figure Appendix C.2 illustrate the age-specific mortality dynamics corresponding to retirement ages of 70 and 75, respectively, providing a detailed comparison with the baseline scenario of retirement at age 65 (Figure 4). As clearly observed, the overall trends in age-structured mortality remain qualitatively consistent across the different retirement age scenarios. Specifically, for the vast majority of age groups—except for individuals aged 100 years and older—the mortality rates exhibit a monotonic decline over time. This pattern persists regardless of whether physician supply is expanded. The exception observed in the 100+ age group likely reflects cumulative survival gains from earlier age groups. As more individuals benefit from adequate medical care and survive into extreme old age, the number of people in the 100+ cohort increases,

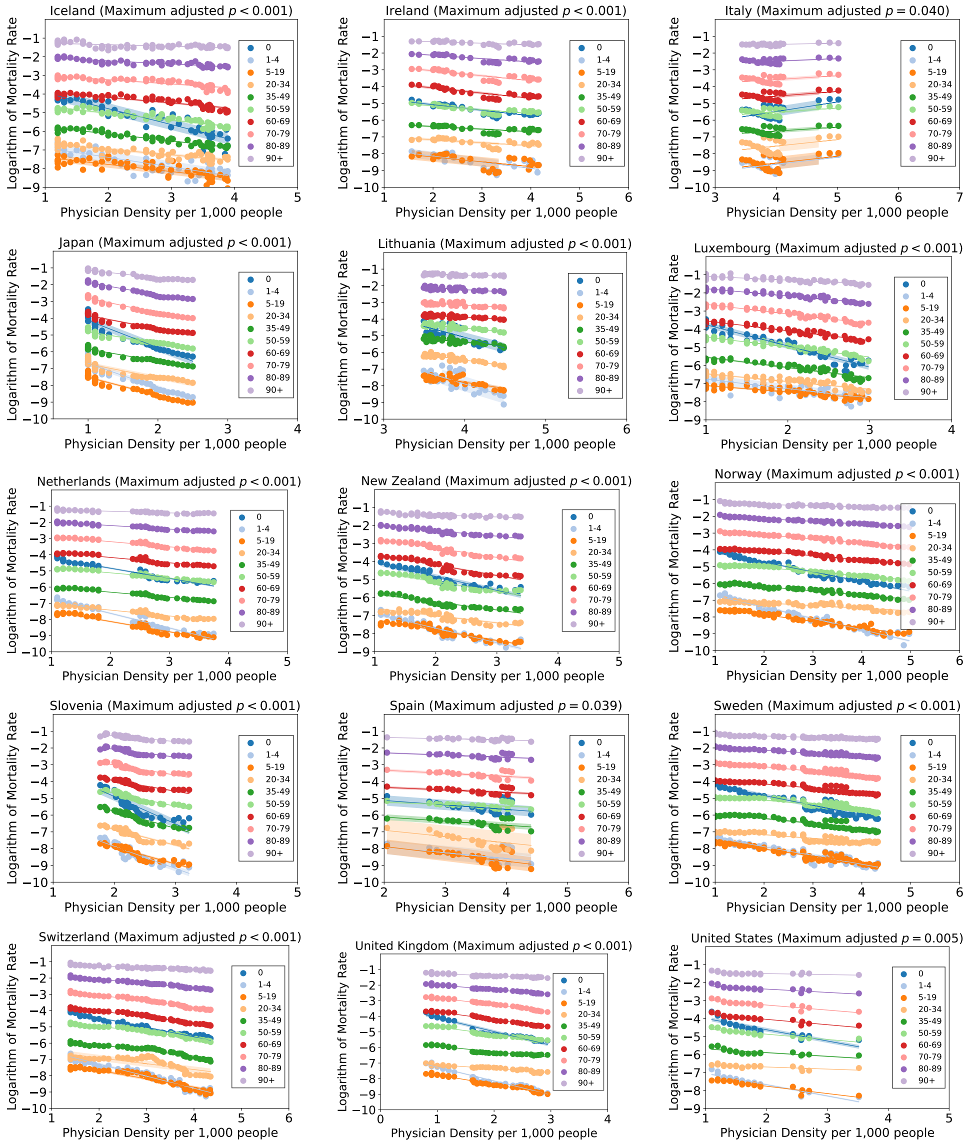


**Figure Appendix B.2:** Linear regression of logarithmic mortality rate against physician density (1960-2022) across all age groups. The solid line represents the mean, with shaded regions indicating 99.7% confidence intervals.

thereby concentrating mortality in this age group. As a result, the mortality rate for the 100+ group increases, not due to worsening health outcomes, but due to the expanded size of the age cohort itself.

We also evaluated the reductions in mortality rates in Table Appendix C.1 and Table Appendix C.2 under retirement ages of 70 and 75, respectively. Compared to Table 1, the reduction percentages were less than 1% relative to the baseline retirement age of $a^* = 65$ (Table 1) in both cases. These results suggest that changes in the retirement age $a^*$ have a negligible impact on age-structured mortality rates. Figure Appendix C.3.(a)–(d) presents the projected population from 2025 to 2065 under three retirement age scenarios (65, 70, and 75). Regardless of the assumed threshold, the overall trends remain consistent. In fact, the projected populations across the three retirement age scenarios do not show statistically significant differences ($p > 0.07$), regardless of the policy applied. These results indicate that the retirement threshold does not critically affect future population projections. Moreover, Figure Appendix C.3.(e)–(l) shows that although varying the retirement age threshold does influence physician density to some extent, the overall physician density continues to increase over time—primarily due to persistently low fertility rates.

We now vary the age-structured fertility rates to examine their impact on future population dynamics. To this end,

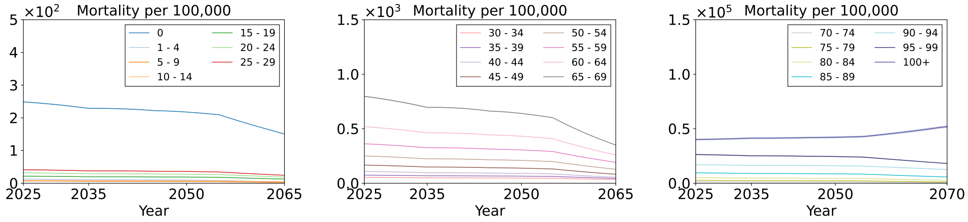


Policy *P1*

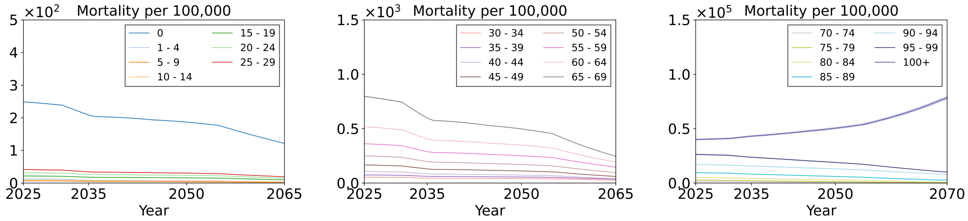


Policy *P2*

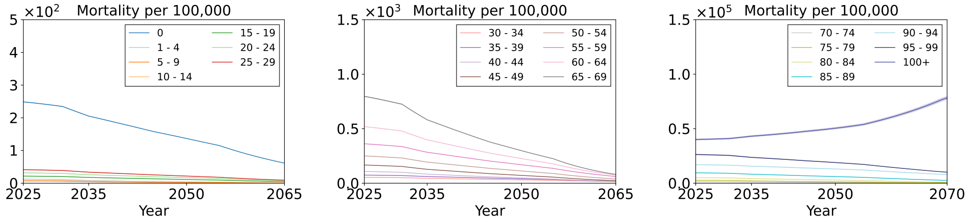


Policy *P3*

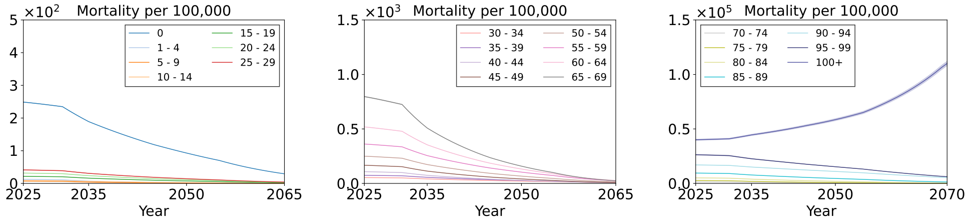


Policy *P4*

**Figure Appendix C.1:** The predicted mortality rates per each age group with **retirement age 70** under (a) *P1*: annual physician supplies 3,058, (b) *P2*: annual physician supplies 3,058 with an increase to 5,058 for the first five years, (c) *P3*: annual physician supplies 5,058, and (d) *P4*: annual physician supplies 7,058. The solid line represents the mean, while the shaded regions indicate the 99.7% confidence intervals.

**Table Appendix C.1:** Expected Mortality Reduction (%) per 100,000 people in **retirement age 70** compared to the original physician supply planning *P1*.

| Age Group | *P2* (%) | *P3* (%) | *P4* (%) | Age Group | *P2* (%) | *P3* (%) | *P4* (%) |
|---|---|---|---|---|---|---|---|
| 0 | 0.02907 | 0.089292 | 0.121325 | 50-54 | 0.031883 | 0.089425 | 0.113566 |
| 01-04 | 0.001594 | 0.003715 | 0.00423 | 55-59 | 0.045577 | 0.13019 | 0.167476 |
| 05-09 | 0.000896 | 0.002087 | 0.002376 | 60-64 | 0.066572 | 0.184349 | 0.232071 |
| 10-14 | 0.001093 | 0.00316 | 0.004099 | 65-69 | 0.103831 | 0.269981 | 0.326364 |
| 15-19 | 0.002601 | 0.007821 | 0.010452 | 70-74 | 0.180501 | 0.479906 | 0.58821 |
| 20-24 | 0.003746 | 0.011396 | 0.015368 | 75-79 | 0.331147 | 0.9628 | 1.254498 |
| 25-29 | 0.004955 | 0.014929 | 0.019981 | 80-84 | 0.585266 | 1.817828 | 2.492045 |
| 30-34 | 0.006105 | 0.019073 | 0.026269 | 85-89 | 0.966207 | 3.192406 | 4.608861 |
| 35-39 | 0.008778 | 0.026768 | 0.036163 | 90-94 | 1.221762 | 4.431698 | 6.968048 |
| 40-44 | 0.013792 | 0.038509 | 0.048754 | 95-99 | 2.220529 | 7.782036 | 11.84527 |
| 45-49 | 0.021474 | 0.059038 | 0.07396 | 100+ | -4.17418 | -20.0326 | -42.5168 |

we consider total fertility rates (TFRs) of 0.72 and 1.05, based on historical age-specific fertility data from South Korea in the years 2023 and 2017, respectively. Table Appendix C.3 presents the age-specific fertility rates (per 1,000 women) for three reference years—2007, 2017, and 2024—corresponding to TFRs of 1.25, 1.07, and 0.72, respectively. In 2007,

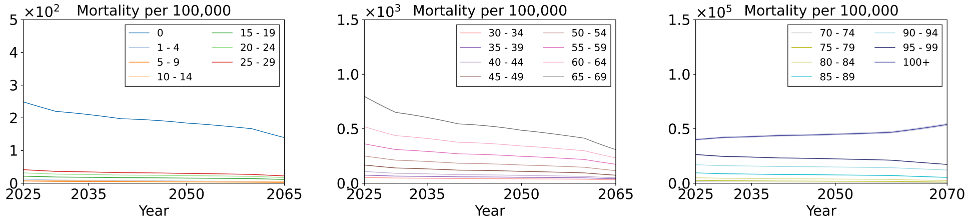


Policy *P1*

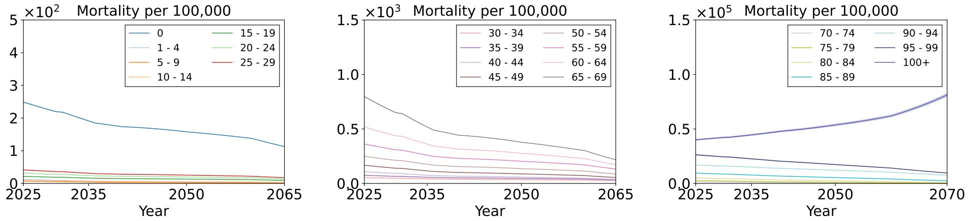


Policy *P2*

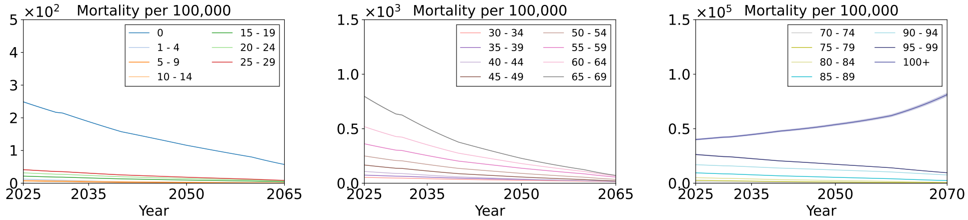


Policy *P3*

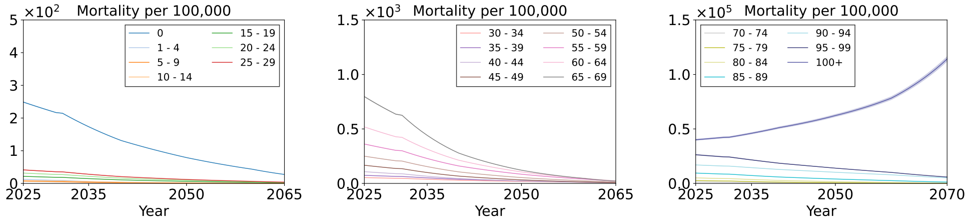


Policy *P4*

**Figure Appendix C.2:** The predicted mortality rates per each age group with **retirement age 75** under (a) *P1*: annual physician supplies 3,058, (b) *P2*: annual physician supplies 3,058 with an increase to 5,058 for the first five years, (c) *P3*: annual physician supplies 5,058, and (d) *P4*: annual physician supplies 7,058. The solid line represents the mean, while the shaded regions indicate the 99.7% confidence intervals.

**Table Appendix C.2:** Expected Mortality Reduction (%) per 100,000 people in **retirement age 75** compared to the original physician supply planning *P1*.

| Age Group | *P2* (%) | *P3* (%) | *P4* (%) | Age Group | *P2* (%) | *P3* (%) | *P4* (%) |
|---|---|---|---|---|---|---|---|
| 0 | 0.026757 | 0.082249 | 0.112036 | 50-54 | 0.028593 | 0.080303 | 0.102225 |
| 01-04 | 0.001346 | 0.003144 | 0.003587 | 55-59 | 0.041095 | 0.117528 | 0.151553 |
| 05-09 | 0.000756 | 0.001766 | 0.002014 | 60-64 | 0.059475 | 0.164924 | 0.20811 |
| 10-14 | 0.000989 | 0.002862 | 0.003722 | 65-69 | 0.090972 | 0.236952 | 0.287076 |
| 15-19 | 0.00238 | 0.007162 | 0.009596 | 70-74 | 0.159261 | 0.424115 | 0.521014 |
| 20-24 | 0.003439 | 0.01047 | 0.014154 | 75-79 | 0.300118 | 0.873542 | 1.140987 |
| 25-29 | 0.004537 | 0.01368 | 0.018354 | 80-84 | 0.540346 | 1.679468 | 2.308162 |
| 30-34 | 0.005646 | 0.017648 | 0.024368 | 85-89 | 0.9069 | 2.997304 | 4.338141 |
| 35-39 | 0.008063 | 0.024608 | 0.033329 | 90-94 | 1.174156 | 4.257443 | 6.710552 |
| 40-44 | 0.012352 | 0.034535 | 0.043827 | 95-99 | 2.115878 | 7.414367 | 11.31384 |
| 45-49 | 0.019143 | 0.052705 | 0.066181 | 100+ | -4.27393 | -20.4589 | -43.4946 |

the highest fertility was observed among women aged 30–34 (102.0), followed closely by those aged 25–29 (95.9), indicating a fertility peak in early adulthood. By 2017, however, fertility in the 25–29 age group had halved to 47.9, while fertility in the 35–39 group nearly doubled to 47.2, reflecting a notable shift toward later childbearing. By 2024,

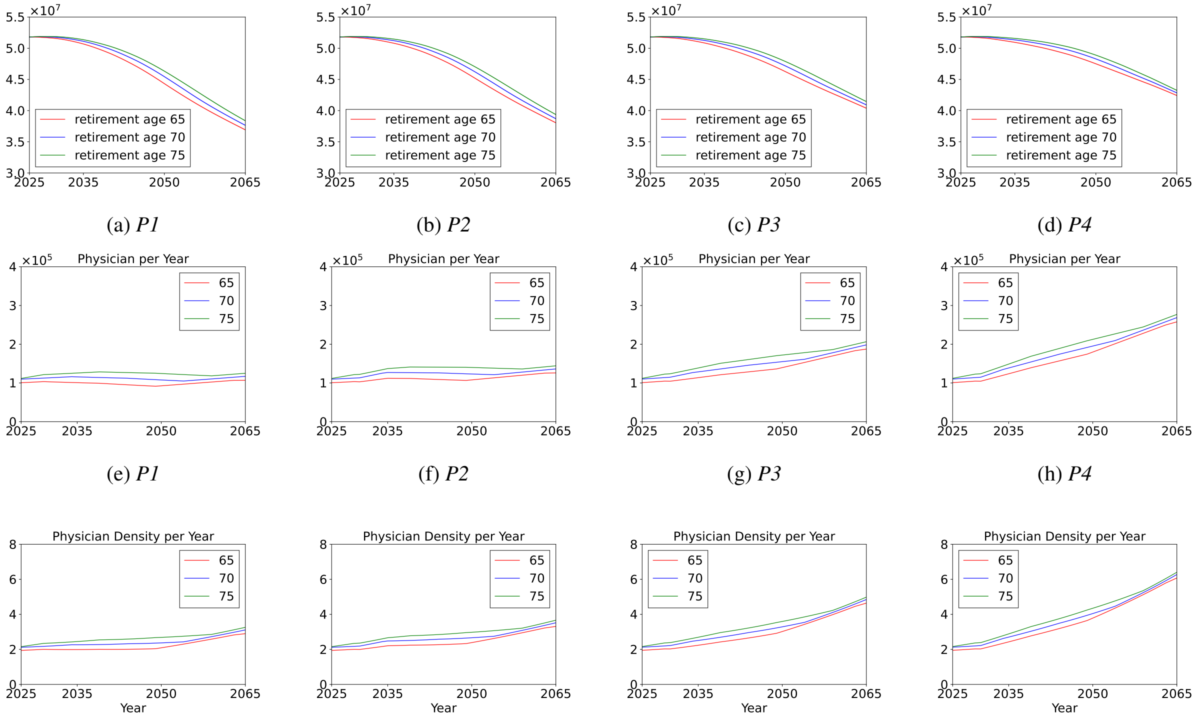


(a) *P1* (b) *P2* (c) *P3* (d) *P4*

(e) *P1* (f) *P2* (g) *P3* (h) *P4*

(i) Policy *P1* (j) Policy *P2* (k) Policy *P3* (l) Policy *P4*

**Figure Appendix C.3:** (a)-(d): Predicted population curves for **different retirement ages** across each supply strategy. Mann-Whitney U test showed no significant differences between different physician retirement threshold.($p \geq 0.185$). (e)-(h): Predicted physicians. (i)-(l): Predicted physician-per-1,000 ratio.

this trend became even more pronounced: fertility rates among younger age groups (15–24) declined sharply, and the rate for the 25–29 cohort dropped to 21.4. Although fertility rates for the 30–34 and 35–39 age groups also declined compared to earlier years, they remained the most significant contributors to national births. These patterns highlight a demographic transition characterized by delayed marriage and childbirth, declining overall fertility, and increased birth concentration among older reproductive age groups.

Figure Appendix C.4 and Figure Appendix C.5 show the age-structured mortality rates under a retirement age of 65 with TFRs of 0.72 and 1.05, respectively. Compared with Figure 4, the age-structured mortality rates under TFRs of 0.72, 1.05, and 1.25 do not exhibit substantial qualitative differences. For each fertility rate, the mortality rates in all age groups—except for the 100+ age group—consistently decline over time. These results suggest that physician density will continue to increase even under the baseline policy scenario *P1*.

Figure Appendix C.6.(a)–(d) present the projected populations under policy scenarios *P1*–*P4* with a retirement age of $a^* = 65$. Because age-structured mortality rates are only marginally affected by changes in fertility, the primary driver of future population trends is the age-specific fertility rate. Furthermore, the number of physicians remains largely unchanged across fertility scenarios, as shown in Figure Appendix C.6.(e)–(h). Consequently, physician densities are directly influenced by fertility-driven changes in population size, as illustrated in Figure Appendix C.6.(i)–(l).

Table Appendix C.4 and Table Appendix C.5 report the expected mortality reductions, analogous to Table 1. Comparing these tables, we find that all age groups—except the 100+ cohort—exhibit less than a 1% improvement when fertility rates are varied.

The results show that in the case of South Korea, future age-structured mortality rates remain largely unaffected by either retirement age or fertility variations. These findings underscore the urgent need for policies focused on improving fertility, as fertility has minimal impact on mortality trends but a direct and substantial influence on demographic structure. Therefore, effective fertility policies are critical to mitigating long-term demographic challenges.

| Age Group | 2007 Rate | 2017 Rate | 2024 Rate |
|---|---|---|---|
| 15–19 | 2.3 | 1.0 | 0.3 |
| 20–24 | 19.7 | 9.6 | 3.8 |
| 25–29 | 95.9 | 47.9 | 21.4 |
| 30–34 | 102.0 | 97.7 | 66.7 |
| 35–39 | 25.9 | 47.2 | 43.0 |
| 40–44 | 3.1 | 6.0 | 7.9 |
| 45–49 | 0.2 | 0.2 | 0.2 |

**Table Appendix C.3:** Age-specific fertility rates per 1,000 women in Korea across years 2007, 2017, and 2024.

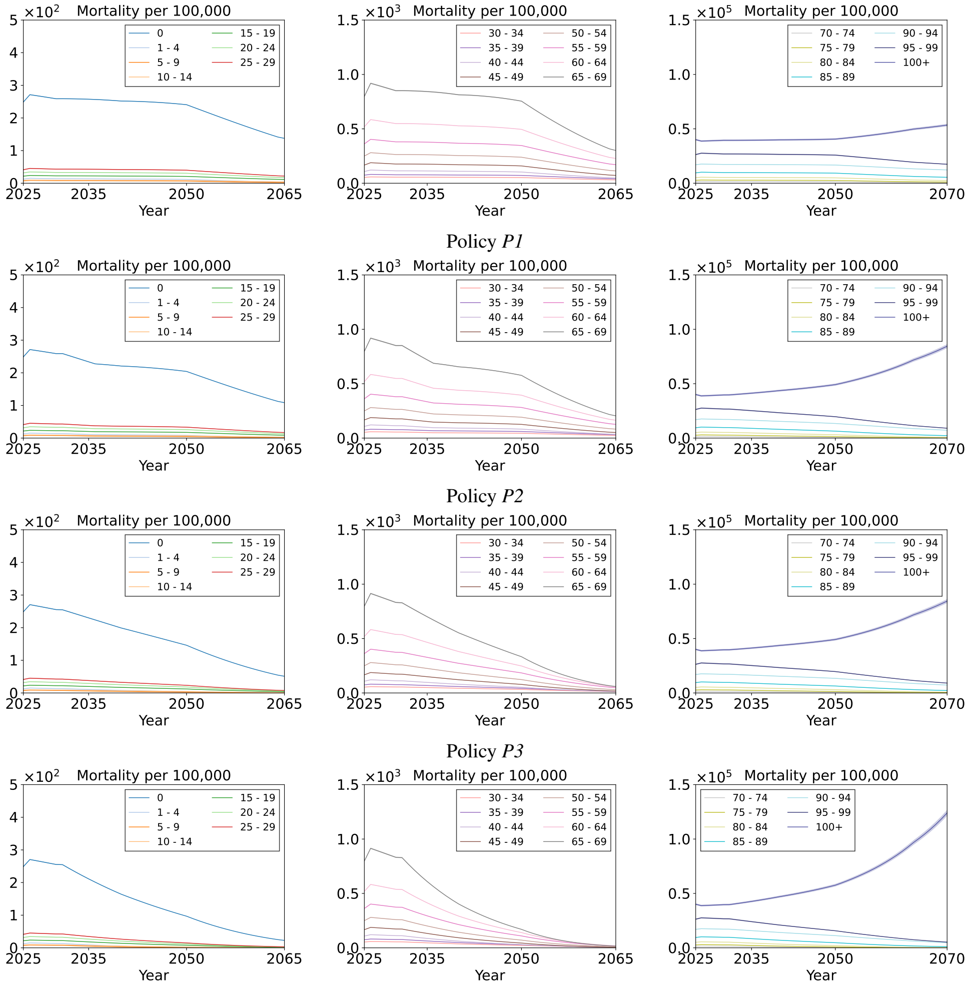


Policy *P1*

Policy *P2*

Policy *P3*

Policy *P4*

**Figure Appendix C.4:** The predicted mortality rates per each age group with retirement age 65 and **TFR 0.72** under (a) *P1*: annual physician supplies 3,058, (b) *P2*: annual physician supplies 3,058 with an increase to 5,058 for the first five years, (c) *P3*: annual physician supplies 5,058, and (d) *P4*: annual physician supplies 7,058. The solid line represents the mean, while the shaded regions indicate the 99.7% confidence intervals.

Furthermore, we consider the following additional scenarios to evaluate the impact of the reduction:

$$P5\text{: } m_t = 3,058 \quad t \leq 6, \qquad \text{otherwise, } m_t = 2,858, \tag{C.1}$$

$$P5\text{: } m_t = 3,058 \quad t \leq 6, \qquad \text{otherwise, } m_t = 2,558, \tag{C.2}$$

$$P7\text{: } m_t = 3,058 \quad t \leq 6, \qquad \text{otherwise, } m_t = 2,058, \tag{C.3}$$

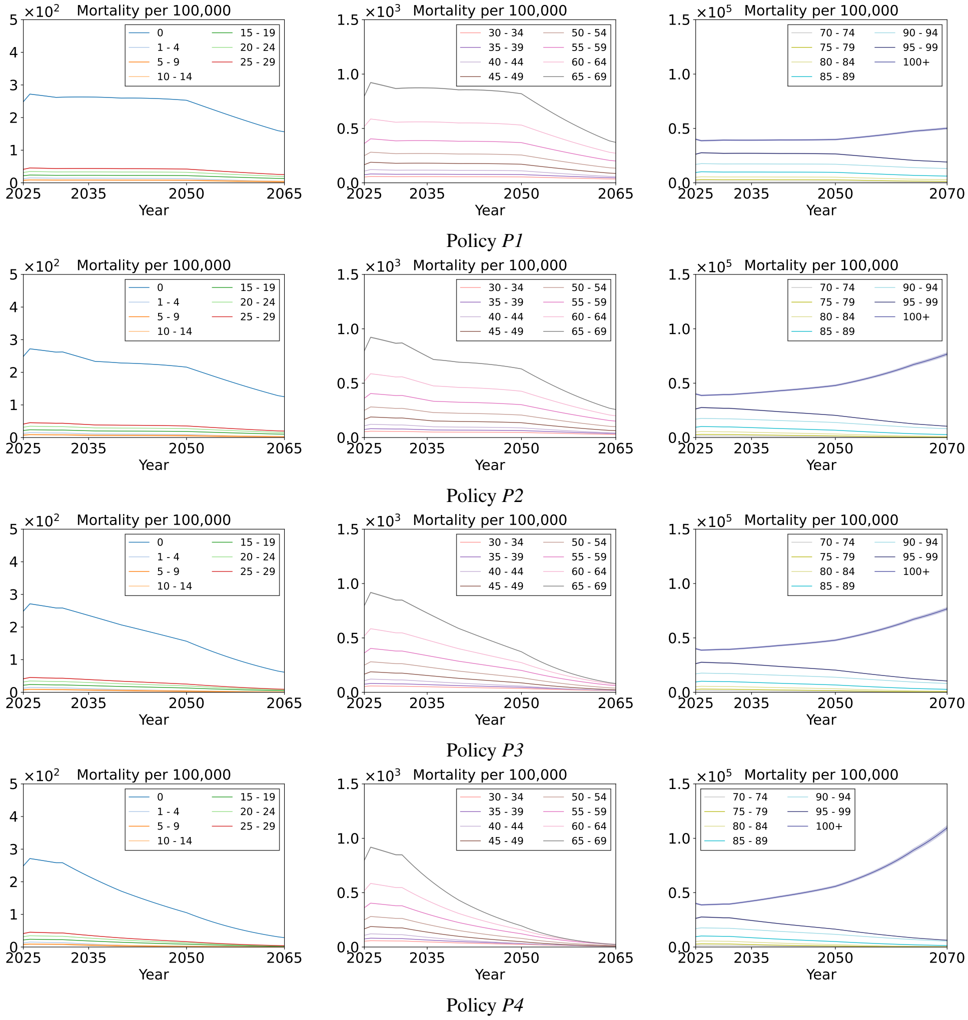


**Figure Appendix C.5:** The predicted mortality rates per each age group with retirement age 65 and **TFR 1.05** under (a) *P1*: annual physician supplies 3,058, (b) *P2*: annual physician supplies 3,058 with an increase to 5,058 for the first five years, (c) *P3*: annual physician supplies 5,058, and (d) *P4*: annual physician supplies 7,058. The solid line represents the mean, while the shaded regions indicate the 99.7% confidence intervals.

**Table Appendix C.4:** Expected Mortality Reduction (%) per 100,000 people in retirement age $a^* = 75$ compared to the original physician supply planning *P1*.

| Age Group | *P2* (%) | *P3* (%) | *P4* (%) | Age Group | *P2* (%) | *P3* (%) | *P4* (%) |
|---|---|---|---|---|---|---|---|
| 0 | 0.028926 | 0.08636 | 0.114765 | 50-54 | 0.030651 | 0.083009 | 0.102953 |
| 01-04 | 0.001415 | 0.003152 | 0.003515 | 55-59 | 0.044129 | 0.121867 | 0.153123 |
| 05-09 | 0.000795 | 0.00177 | 0.001974 | 60-64 | 0.063677 | 0.170114 | 0.209134 |
| 10-14 | 0.001063 | 0.002974 | 0.003768 | 65-69 | 0.096793 | 0.241845 | 0.285596 |
| 15-19 | 0.002567 | 0.007493 | 0.009789 | 70-74 | 0.169833 | 0.434481 | 0.520114 |
| 20-24 | 0.003714 | 0.010975 | 0.014471 | 75-79 | 0.322813 | 0.908541 | 1.156496 |
| 25-29 | 0.004896 | 0.014315 | 0.018729 | 80-84 | 0.584722 | 1.766822 | 2.369726 |
| 30-34 | 0.006112 | 0.018585 | 0.025047 | 85-89 | 0.986706 | 3.187661 | 4.513383 |
| 35-39 | 0.008711 | 0.025805 | 0.034091 | 90-94 | 1.287496 | 4.604156 | 7.139813 |
| 40-44 | 0.013235 | 0.035672 | 0.044105 | 95-99 | 2.313559 | 7.968574 | 11.93463 |
| 45-49 | 0.020481 | 0.054297 | 0.066426 | 100+ | -4.78773 | -23.3054 | -50.2554 |

*P5–P6* in Eqs. Eq. (C.1)–Eq. (C.3) represent reductions in annual physician supply compared to *P1*. Figure Appendix C.7 presents mortality rates under different physician supply scenarios: *P1* (3,058 physicians annually), *P5* (2,858), *P6* (2,558), and *P7* (2,058), as defined in Eqs. Eq. (C.1)–Eq. (C.3). These results are statistically valid with probability

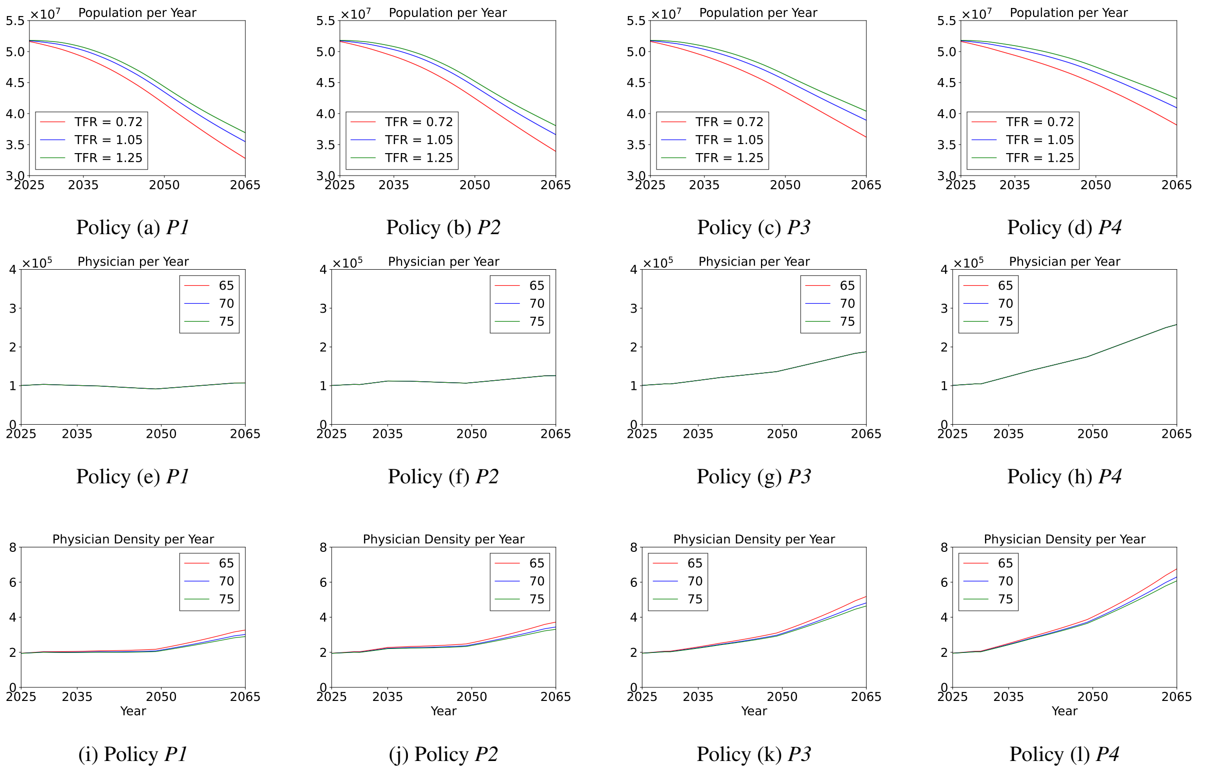


**Figure Appendix C.6:** Projections for different TFRs across different policies. (a)-(d): Predicted population curves under each supply strategy. (e)-(h): Predicted physicians. (i)-(l): Predicted physician-per-1,000 ratio.

**Table Appendix C.5:** Expected Mortality Reduction (%) per 100,000 people in retirement age $a^* = 75$ compared to the original physician supply planning *P1*.

| Age Group | *P2* (%) | *P3* (%) | *P4* (%) | Age Group | *P2* (%) | *P3* (%) | *P4* (%) |
|---|---|---|---|---|---|---|---|
| 0 | 0.031283 | 0.094803 | 0.127671 | 50-54 | 0.034688 | 0.095752 | 0.120456 |
| 01-04 | 0.001778 | 0.004064 | 0.004589 | 55-59 | 0.049474 | 0.13915 | 0.177325 |
| 05-09 | 0.001 | 0.002284 | 0.002578 | 60-64 | 0.072544 | 0.197646 | 0.246463 |
| 10-14 | 0.001185 | 0.003373 | 0.004335 | 65-69 | 0.114082 | 0.291414 | 0.348988 |
| 15-19 | 0.002805 | 0.008319 | 0.011017 | 70-74 | 0.197733 | 0.51673 | 0.62739 |
| 20-24 | 0.004036 | 0.01211 | 0.016184 | 75-79 | 0.358681 | 1.027322 | 1.326133 |
| 25-29 | 0.005344 | 0.015878 | 0.021058 | 80-84 | 0.629002 | 1.92817 | 2.620208 |
| 30-34 | 0.006557 | 0.02022 | 0.027608 | 85-89 | 1.031234 | 3.369131 | 4.825842 |
| 35-39 | 0.009454 | 0.028438 | 0.038076 | 90-94 | 1.291156 | 4.645281 | 7.261628 |
| 40-44 | 0.015013 | 0.041252 | 0.051734 | 95-99 | 2.355045 | 8.176734 | 12.36338 |
| 45-49 | 0.023423 | 0.063344 | 0.078604 | 100+ | -4.29631 | -20.6682 | -44.0873 |

0.972, under Assumptions 1–5. Figure Appendix C.7(d) shows a significant increase in mortality rates for several age groups (e.g., the age-0 group) under *P7* from 2035 to 2050. Conversely, Figure Appendix C.7(a)–(c) indicate that the trends under *P1*, *P5*, and *P6* exhibit nearly flat, slightly decreasing slopes. These findings suggest that maintaining at least 2,858 physicians per year (*P5*) ensures consistent reductions in mortality rates, making it a minimally rational planning choice that balances improved health outcomes with financial sustainability.

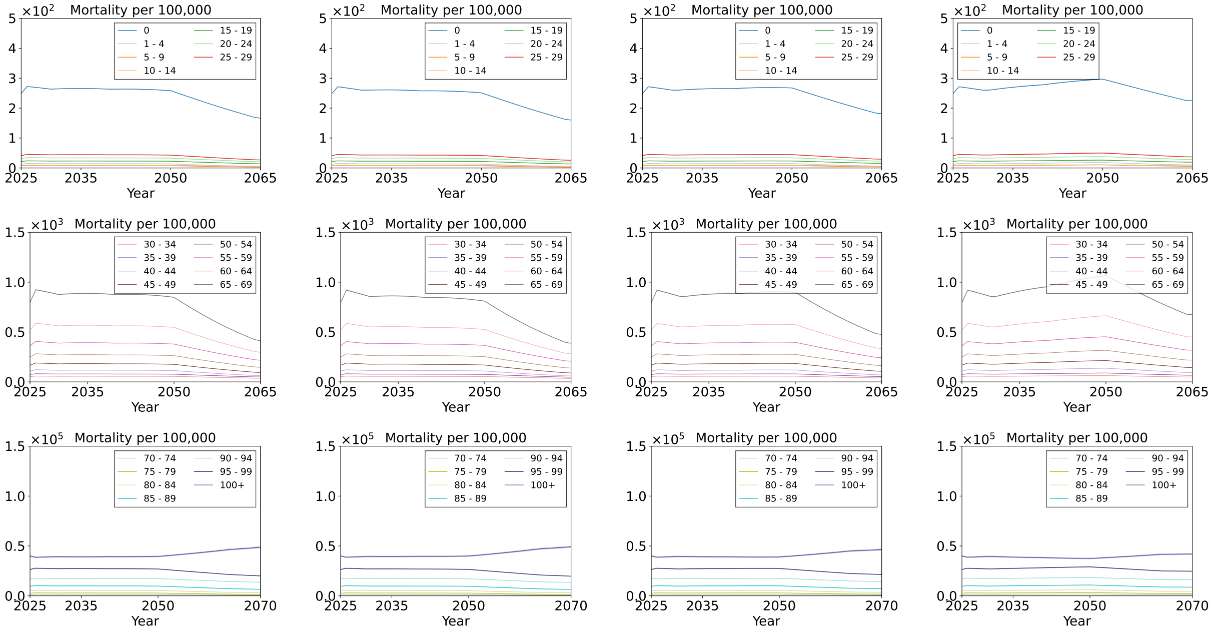


(a) Policy *P1* (b) Policy *P5* (c) Policy *P6* (d) Policy *P7*

**Figure Appendix C.7:** The predicted mortality rates per each age group under (a) *P1*: annual physician supplies 3,058, (b) *P5*: annual physician supplies 2,858, (c) *P6*: annual physician supplies 2,558, and (d) *P7*: annual physician supplies 2,058. The solid line represents the mean, while the shaded regions indicate the 99.7% confidence intervals.
